# Image-Based Consensus Molecular Subtyping in Rectal Cancer Biopsies and Response to Neoadjuvant Chemoradiotherapy

**DOI:** 10.1101/2023.10.26.23297521

**Authors:** Maxime W Lafarge, Enric Domingo, Korsuk Sirinukunwattana, Ruby Wood, Leslie Samuel, Graeme Murray, Susan D Richman, Andrew Blake, David Sebag-Montefiore, Simon Gollins, Eckhard Klieser, Daniel Neureiter, Florian Huemer, Richard Greil, Philip Dunne, Philip Quirke, Lukas Weiss, Jens Rittscher, Tim Maughan, Viktor H Koelzer

## Abstract

The development of deep learning (DL) models to predict the consensus molecular subtypes (CMS) from histopathology images (imCMS) is a promising and cost-effective strategy to support patient stratification. Here, we investigate whether imCMS calls generated from whole slide histopathology images (WSIs) of rectal cancer (RC) pre-treatment biopsies are associated with pathological complete response (pCR) to neoadjuvant long course chemoradiotherapy (LCRT) with single agent fluoropyrimidine.

DL models were trained to classify WSIs of colorectal cancers stained with hematoxylin and eosin into one of the four CMS classes using a multi-centric dataset of resection and biopsy specimens (n=1057 WSIs) with paired transcriptional data. Classifiers were tested on a held out RC biopsy cohort (ARISTOTLE) and correlated with pCR to LCRT in an independent dataset merging two RC cohorts (ARISTOTLE, n=114 and SALZBURG, n=55 patients).

DL models predicted CMS with high classification performance in multiple comparative analyses. In the independent cohorts (ARISTOTLE, SALZBURG), cases with WSIs classified as imCMS1 had a significantly higher likelihood of achieving pCR (OR=2.69, 95%CI 1.01-7.17, p=0.048). Conversely, imCMS4 was associated with lack of pCR (OR=0.25, 95%CI 0.07-0.88, p=0.031). Classification maps demonstrated pathologist-interpretable associations with high stromal content in imCMS4 cases, associated with poor outcome. No significant association was found in imCMS2 or imCMS3.

imCMS classification of pre-treatment biopsies is a fast and inexpensive solution to identify patient groups that could benefit from neoadjuvant LCRT. The significant associations between imCMS1/imCMS4 with pCR suggest the existence of predictive morphological features that could enhance standard pathological assessment.

## INTRODUCTION

Important progress has been made in the treatment of high-risk rectal cancer (RC) patients in the past decades [1]. Prospective clinical trials of neoadjuvant chemoradiotherapy have shown that approximately 15% of patients can reach pathological complete response (pCR) by neoadjuvant chemoradiotherapy (CRT) [2] and that response rates can be further increased to about 30% pCR by total neoadjuvant treatment (TNT) [3–6]. Organ sparing approaches may be realistic for patients with pCR, and can have major impact on long-term quality after definite treatment. However, intensified neoadjuvant treatment is associated with increased frequency and severity of adverse events that can present during or after CRT [7]. At the same time, responses are heterogeneous with 30-40% of patients presenting with tumour regression of different grades while 7-30% of patients classify as non-responders [8, 9]. Interpretable predictive biomarkers to guide personalized treatment are therefore of utmost importance to further improve patient selection for intensified treatment regimens and treatment outcomes [10]. The setting for the identification of predictive markers in the clinical pathway of RC patients remains highly challenging: Turnaround time needs to be short and only scarce material from diagnostic biopsies is available for study, limiting the utility of molecular and functional analysis methods.

The Consensus Molecular Subtypes (CMS) define four distinct subtypes of colorectal cancer (CRC) by common patterns of gene expression [11]. The prognostic value of CMS classes has been reported in several studies [12, 13], yet their association with treatment outcome is an open research topic [11]. Recently, Domingo et al. [14] have described an association between pCR to neoadjuvant chemoradiotherapy and transcriptional CMS signatures in diagnostic RC biopsies, suggesting that molecular classification may also be used as a predictive biomarker for treatment stratification. Overall, early profiling of CMS classification in the clinical pathway promises to be highly relevant for personalized treatment decisions [15]. Limitations of transcriptional CMS classification include a high cost and time requirement for RNA sequencing, a high failure rate due to the small amount of tumour material available and difficulty to standardize single samples [13, 16]. In addition, even successful transcriptome generation shows higher frequencies of unclassified cases in biopsies than resections [13, 17].

Machine learning has been shown to be a promising alternative solution for predicting molecular signatures across multiple cancer types from diagnostic histopathology sections [18]. In particular, Sirinukunwattana et al. [13] showed that deep learning models can predict image-based CMS (imCMS) classes that match transcriptional CMS calls directly from hematoxylin-and-eosin (H&E) stained histopathology whole slide images (WSIs) of CRC tumour specimens, using three independent cohorts. This prior work also highlighted the potential of imCMS to stratify patients for whom transcriptional CMS failed. Yet, evidence of the generalization of imCMS on biopsy cohorts was limited by the need for domain-adversarial training and fine-tuning, requiring labeled data from the target cohort, which is a shortcoming for the implementation of imCMS in the clinic.

Here, we set out to show whether imCMS classifiers can be trained without domain adaptation and perform reproducibly in independent pre-treatment biopsy datasets. By investigation of real-world cohorts sourced from the Medical Research Council (MRC) and Cancer Research UK (CRUK) Stratification in Colorectal cancer (S:CORT), we estimated via simulation experiments how many cancer biopsies are sufficient to achieve near-optimal imCMS classification performance. We then assessed whether imCMS classification is associated with pCR to neoadjuvant long course chemoradiotherapy (LCRT) with single agent fluoropyrimidine treatment in two independent held out diagnostic biopsy cohorts, comprising a total of 169 advanced RC patients with outcome data.

## RESULTS

### Effect of multi-cohort training on imCMS performance

With the goal of developing a generic imCMS classifier that can perform well on both resection and biopsy specimens (without the need for a domain adaptation procedure as required in imCMSv1 [13]), we first assessed the effect of combining resection and biopsy cohorts for training the models, while keeping independent resection and biopsy cohorts held out for evaluation purposes. We designed five experiments with different combinations of cohorts (Figure 1B). For each experiment, the development datasets were split into five bins (with stratified cohorts and classes at the patient level) to build five folds of training/validation splits by using each bin once as an internal validation set (for model selection purposes), while the remaining four bins are used for training. Classification performances were separately measured for each trained model using the corresponding held out test sets, as well as for the ensemble model that combines the output of the five models (Figure 1C).

**Figure 1.**
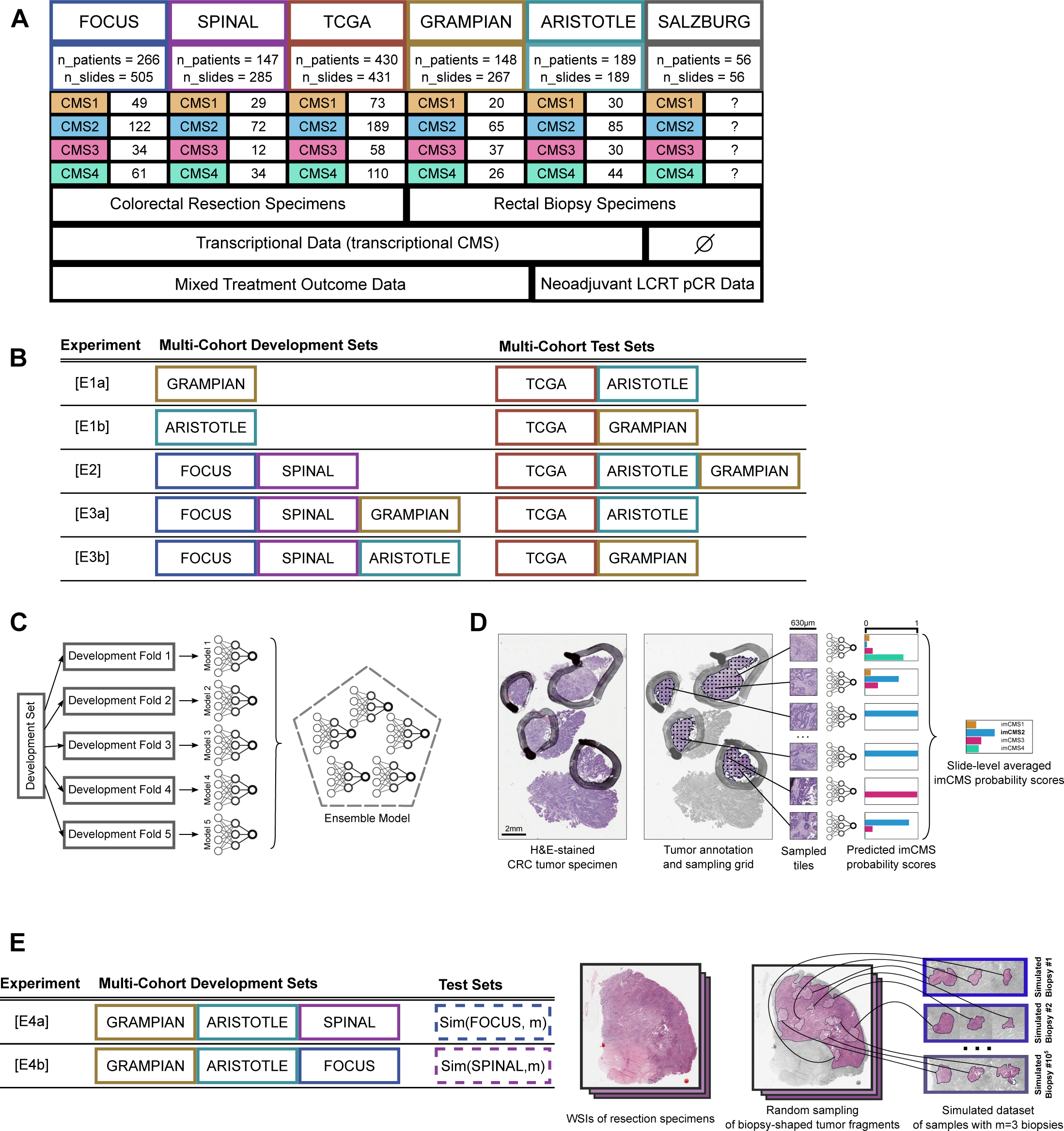
Study Flow Figure. (A) Description of the image dataset used in this study. (B) Table of the cohort-level partitions of the dataset for training and validation of the models compared in this study. (C) Illustration of the 5-fold experimental setup and ensembling approach investigated in this study. (D) Flowchart of the 3-stage imCMS classification pipeline investigated in this study. (E) Table of the cohort-level partitions of the dataset used for training and validation of biopsy simulations (left), and illustration of the biopsy sampling simulation protocol investigated in this study.

The macro-average area under the receiver operating characteristic curves (AUROC) of the trained models are reported in (Figure 2), the detailed ROC curves, and confusion matrices of the best performing trained models are reported in (Supplemental Figure S2). For each test set, we achieved the best performance when both resection and biopsy images were used jointly for training. The ensemble model resulting from the experiment [E3a] (TCGA: macro-average AUROC .813; ARISTOTLE: macro-average AUROC .798) was used for subsequent analysis of outcome association. To ensure a valid assessment of performance, all the datasets in this study were curated by excluding WSIs with poor overall quality (e.g., tissue folding, out-of-focus images), and image processing was restricted to regions of tumor and microenvironment that were delineated by a board-certified pathologist specialised in gastrointestinal pathology. The exclusion criteria used in this study are listed in (Supplemental Figure S1). Robustness to heterogeneity of image appearance and to staining variability was addressed by the application of standard data augmentation policies during training. Details on the training procedure are provided in Supplementary Material and Methods.

**Figure 2.**
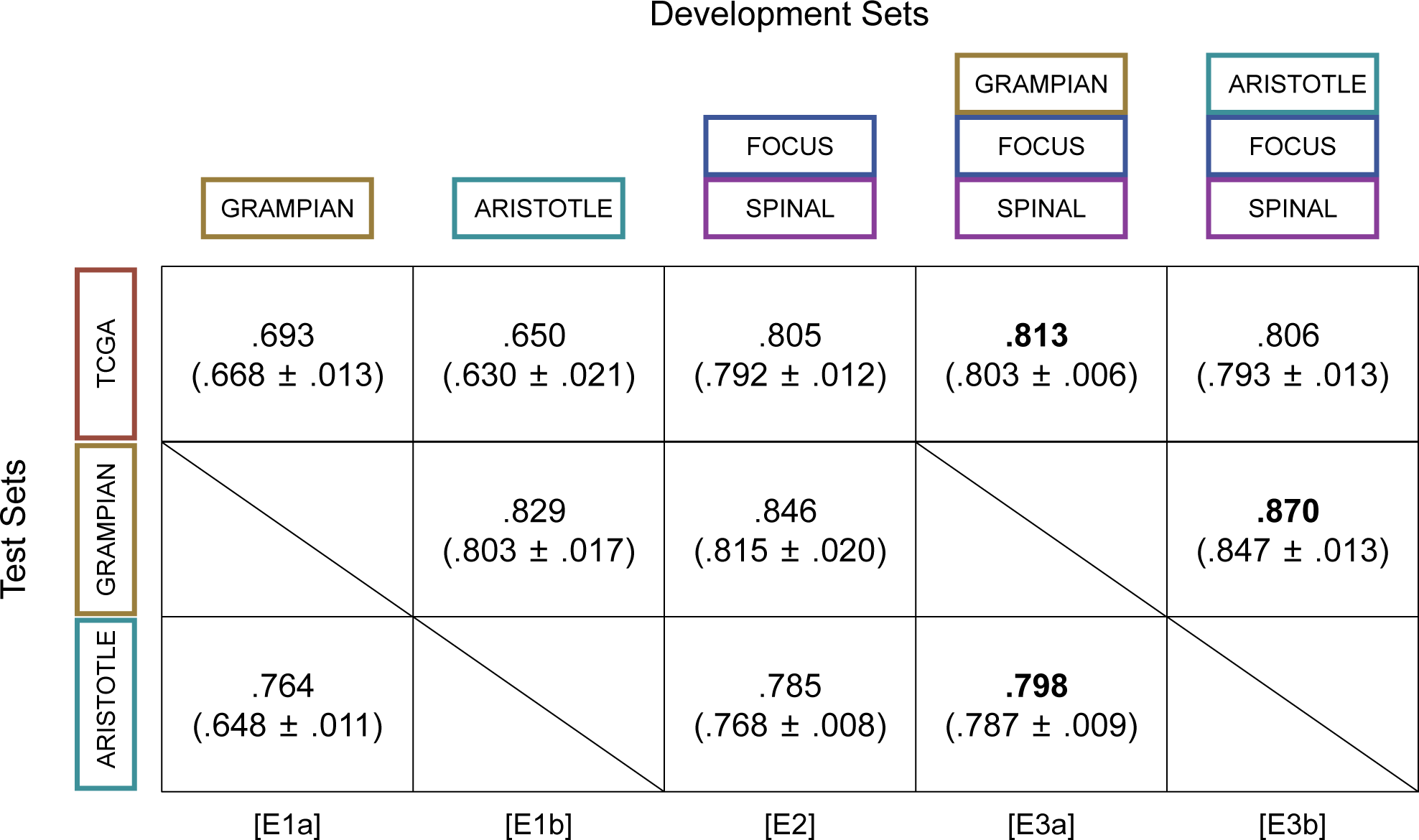
Classification performances of imCMS models trained and evaluated using data from different combinations of cohort. Numbers are macro-averages of the AUROC for ensemble models of five models trained and validated with five different folds of the same development sets. Each model of an ensemble model was evaluated separately: the mean and standard deviation of the corresponding macro-average AUROCs is reported in parenthesis. The ensemble model resulting of the experiment [E3a] was used for subsequent analysis of association between imCMS and pCR to neoadjuvant LCRT.

### Association between imCMS and pCR to neoadjuvant LCRT

To investigate the association between imCMS calls and pCR to neoadjuvant LCRT with single agent fluoropyrimidine, we identified two RC cohorts treated with the same treatment regimen (pelvic irradiation 45-50.4Gy in 25 fractions over 5 weeks, combined with single agent Capecitabine on treatment days) (Supplementary Material and Methods). We applied the imCMS ensemble model resulting from experiment [E3a] to WSIs of all pre-treatment biopsies of the ARISTOTLE and SALZBURG cohorts with available pCR and pre-treatment T/N stage data. imCMS calls at the case level were defined as the majority vote of the calls of the five different trained models that constitute the ensemble model. Cases with undecided majority were classified as “mixed” (n=2). The analysis of imCMS calls with clinicopathological data and treatment outcomes was conducted with 114 patients of the ARISTOTLE cohort including 24 patients with pCR to neoadjuvant LCRT, and 55 patients of the SALZBURG cohort with 6 patients with pCR to neoadjuvant LCRT. Patients were split into groups based on their predicted imCMS class. Odds ratios for pCR were calculated separately for each group and adjusted by T-stage, N-stage and cohort. Significant associations (p-value<0.05) were found for both imCMS4 with lack of pCR (OR 0.25, 95%CI 0.07-0.88, p=0.03) and imCMS1 with pCR (OR 2.69, 95%CI 1.01-7.17, p=0.048) (Figure 3), suggesting that imCMS calls generated from pre-operative biopsy material associate with response of the primary tumour to combined chemoradiotherapy.

**Figure 3.**
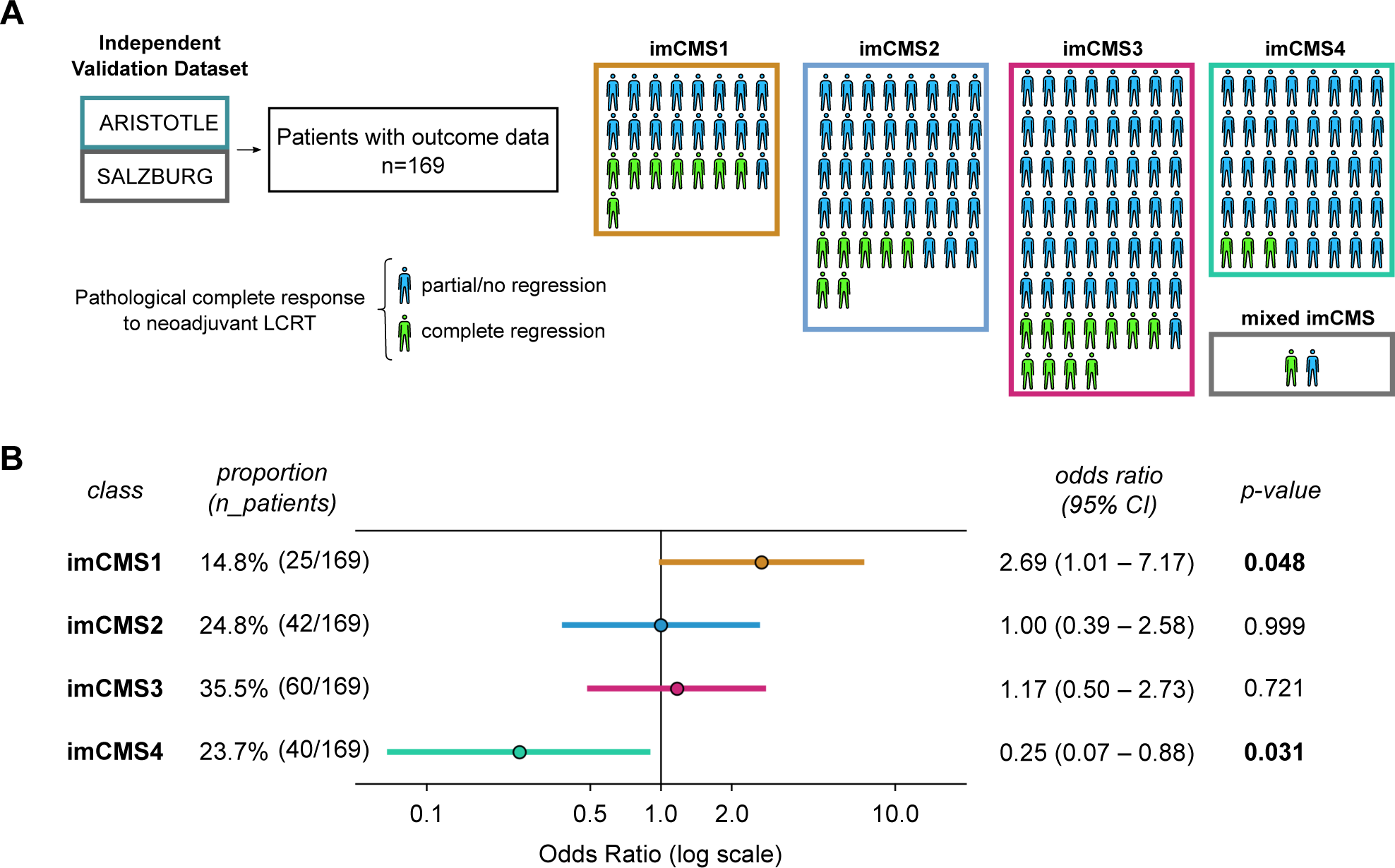
(A) Illustration of the analysis of association between imCMS groups resulting from the application of an ensemble model (experiment [E3a]) and outcome in the independent holdout cohorts ARISTOTLE and SALZBURG. (B) Comparison of odds ratio of pCR outcome by neoadjuvant LCRT between different groups of patients of the ARISTOTLE and SALZBURG cohorts based on imCMS calls derived from pre-operative biopsy images. Patients with biopsies classified as imCMS1 were significantly more likely to achieve pCR by LCRT (OR 2.69; 95%CI: 1.01-7.17, p=0.048) than all other imCMS groups, whereas patients with imCMS4 calls rarely achieved pCR (OR 0.25; 95%CI: 0.07-0.88, p=0.031). Two patients had mixed imCMS classification calls and were not shown in the odds ratio comparison.

### Simulation: Effect of biopsy sampling on imCMS classification

To assess how reliable biopsy-based imCMS predictions are in comparison to resection-based imCMS predictions in a control scenario, we estimated the change in classification performance of imCMS as a function of the number of biopsy fragments in a sample. It is theoretically possible to conduct an experiment in which we would generate a dataset with many tissue fragments biopsied from each patient’s tumour site along with paired resection specimens and associated CMS calls, then comparing imCMS classification performance when combining different numbers of fragments. However, since repeated sampling cannot be ethically justified, we argue that such an experimental protocol can be approximated via the generation of virtual biopsy fragments randomly sampled from existing resection images.

We generated 26 simulations of biopsy datasets, each with a specified number *m* of biopsy fragments and sourced from either of the two datasets of resection specimens, FOCUS or SPINAL. For every WSI in a given resection dataset, we collected 10, 000 subsets of *m* non-overlapping biopsy-fragment-shaped images randomly cropped from the annotated tumor regions in the resection. Cropping was performed by sourcing real shapes of tumour biopsy fragments (Figure 4A-B) that had been manually annotated in the GRAMPIAN and ARISTOTLE cohorts (total of 1580 possible shapes). Examples of simulated biopsy samples from a source resection specimen are illustrated in (Figure 4C-D). Thus, each random subset of *m* fragments simulates a single random biopsy sampling event that an endoscopist could potentially perform, including possibly both superficial and deep samples. Then, all the random subsets generated across all the WSIs of a resection dataset were concatenated to represent a set of random biopsy sampling events (Figure 4C-D). By restricting the cropping procedure to annotated regions of tumor and microenvironment we aimed at having simulated biopsies that are a good representation of real-world biopsies. We designed these simulated datasets to approximate the characteristics of actual biopsy datasets under the assumption that, (a) sampling locations are uniformly distributed in a tumour region, (b) the shape of the biopsies is random and independent of the sampling location, (c) sampling locations of consecutive biopsy fragments are random and non-overlapping. Consequently, we argue that the distribution of images of such a simulated biopsy from a single resection image closely approximates the distribution of images of actual random biopsy samples excised from the same tumour specimen.

**Figure 4.**
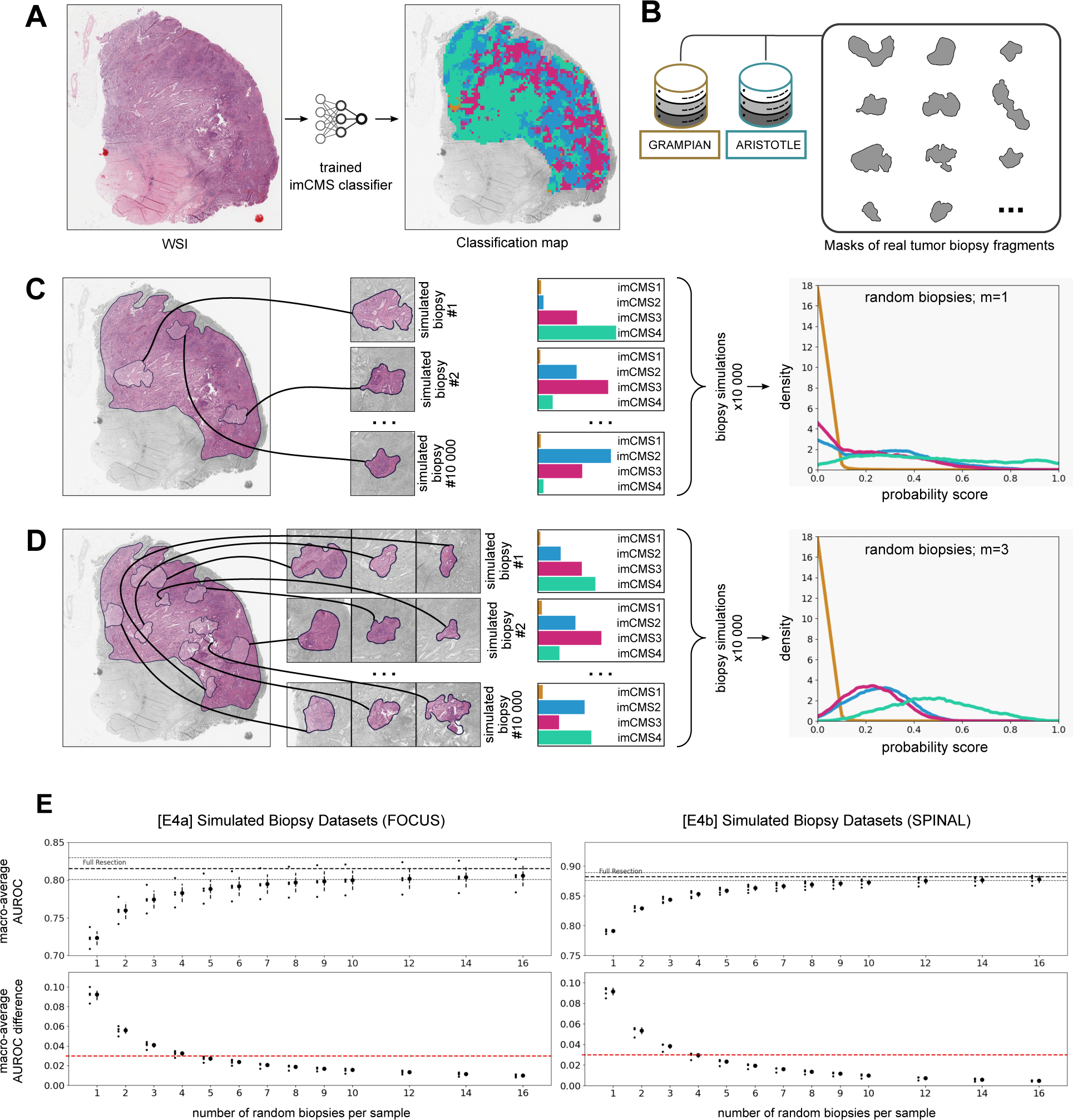
Biopsy Simulation Experiments. (A) Example of a WSI with heterogeneous tile-level imCMS classification. (B) The shapes of simulated biopsy fragments are randomly sampled from annotations of biopsies in the GRAMPIAN and ARISTOTLE cohorts. (C) Illustration of a biopsy simulation procedure based on a source imaged resection specimen. Biopsy samples are randomly generated by sampling m=1 biopsy fragments at random in the annotated tumor region of the resection specimen. This random process is repeated 10 000 times for each WSI of a resection dataset to form a *simulated biopsy dataset*. For each generated biopsy sample, imCMS is applied. The resulting distributions of class probability over a *simulated biopsy dataset* are shown on the right. (D) same as (C) but with m=3 biopsy fragments. The spatial heterogeneity of tile-level predictions (shown in (A)) explains the different distributions obtained for each imCMS class across a simulated biopsy dataset. (E) Classification performance of trained imCMS models on simulated biopsy datasets as a function of the number of tumour biopsy fragments per sample. Dots with lines indicate the mean and standard deviation of the macro-average AUROC obtained by five models for each simulated dataset: [E4a] (left); [E4b] (right). The red dotted line indicates a 3% difference score with the macro-average AUROC obtained on corresponding fully imaged resection specimens.

To assess the performance of imCMS in the simulated biopsy datasets, we made sure to evaluate models on data not seen during training by using the training and test partitions described in (Figure 1E). Each trained imCMS model resulting from the training procedures [E4a] and [E4b] was applied to all its respective simulated test sets. As a result, we observed a rapid increase of classification performances when the number of fragments in a biopsy sample increases until convergence to the performance level that the models achieved when using the resection data without sampling (Figure 4E). For both test conditions, we report a AUROC difference lower than 3% of the AUROC achieved with the original resection data when the number of simulated tumour biopsy fragments in a sample is above five, suggesting that reliable imCMS classification can be achieved at diagnosis in a large fraction of cases [19].

### Stability of distribution of cell types between resection and biopsy samples

As endoscopists excise biopsies in specific targeted regions of tumours, this procedure is subject to variance between operators as well as patient-specific factors. This can potentially induce a variation of morphological information in images of biopsies in comparison to resection specimens. This may particularly affect heterogeneously distributed factors in the tumour microenvironment such as cancer, immune and stromal cell populations. We therefore investigated whether there are systematic differences in microenvironment composition between biopsies and resection specimens of CRC specimens. Specifically, we confirmed the absence of confounding factors with real-world biopsies and investigated whether systematic differences of cell distribution exist between biopsy samples and resection specimens by deconvolution of stromal, immune and tumour-related signatures from transcriptomic data in 529 biopsies and 565 resections from four S:CORT cohorts (FOCUS, SPINAL, GRAMPIAN and ARISTOTLE). First, we generated absolute abundance estimates for key immune and stromal cell types with two different tools (MCPcounter [20] and xCell [21]) using bulk transcriptomic data. With both methods, levels of key immune lineages (T-cells including CD8+ and differentiated cytotoxic T-cell sub-populations, B-cells, NK-cells, monocytes, myeloid dendritic cells and neutrophils) as well as endothelial and stromal cell populations in biopsies and resections were statistically similar across CMS subtypes (all p-values *>* 0.25, ANOVA) (Figure 5). This result suggests that biopsies may be representative of the broad abundance of cell types in the tumour microenvironment compared to resection specimens, according to their transcriptomic CMS classes.

**Figure 5.**
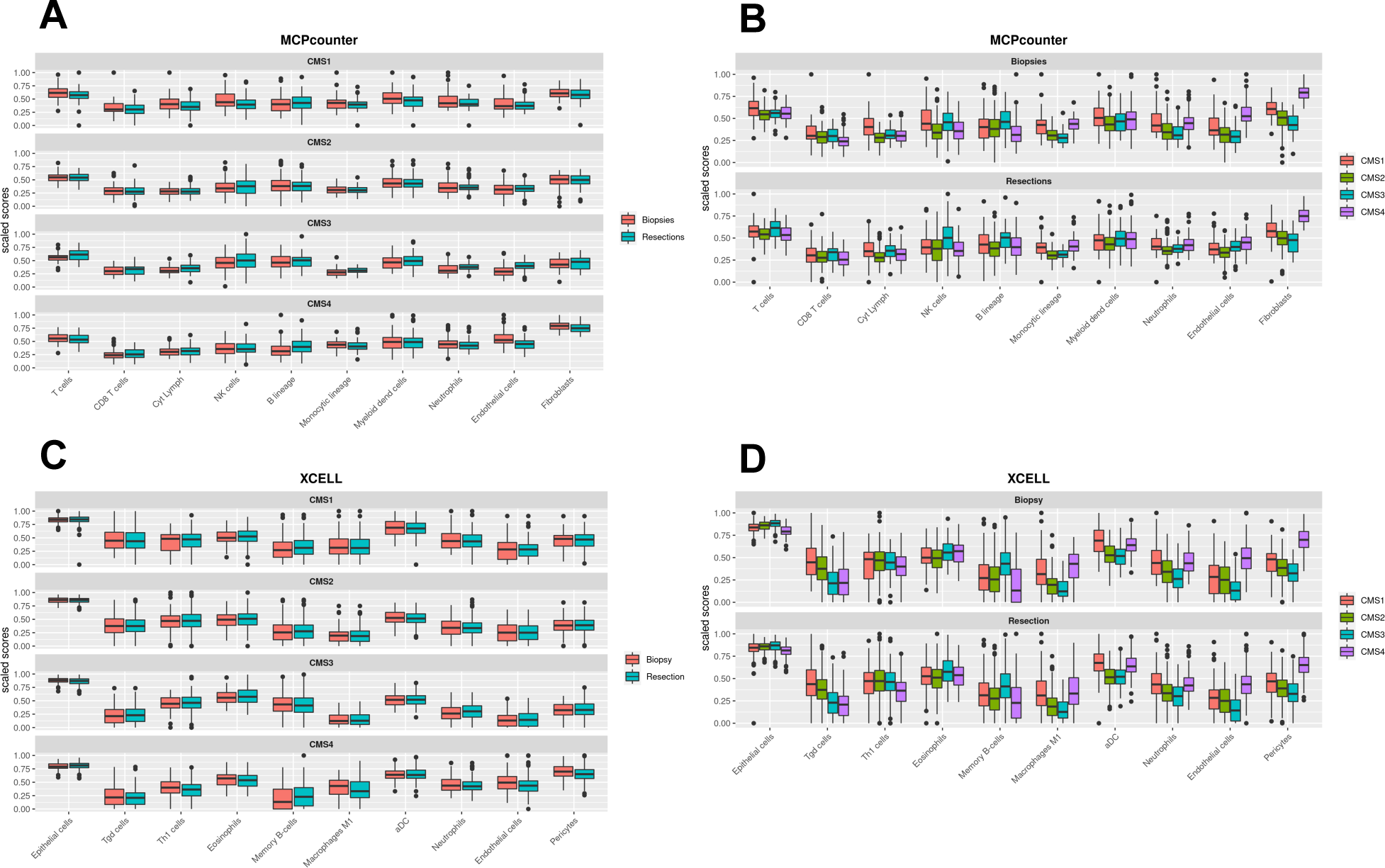
Comparison of measures of abundance scores for ten representative cell types between 529 biopsy and 565 resection primary CRC samples of the cohorts used in this study (FOCUS, SPINAL, GRAMPIAN and ARISTOTLE) using the transcriptome-based MCP-counter (A,B) and xCell tools (C,D). Cell enrichment scores were measured both by CMS subgroup (B,D) and sample type (A,C). Cell type distributions by CMS-class do not differ between biopsy and resection samples, indicating that transcriptional CMS calls derived from biopsy samples robustly capture the underlying biology of CRC shown on the actual tissue. Distributions in panels A and C are similar (p*>* 0.25, ANOVA), whereas all p-values in panels B and D are significant (p*<* 0.05).

### Tissue microenvironment patterns related to CMS classes

CMS classification is driven by microenvironment and tumour-related factors. To investigate consistency of imCMS classification in RC biopsy material with underlying biological patterns, we visualized image tiles with the highest predicted probability score for each imCMS class in biopsy material (Figure 6). This approach provides an overall idea of the morphology associated with each CMS class in absence of an established definition of transcriptional CMS at the scale of image patches. In agreement with prior studies, we found a consistent pattern of high stromal content and dissociative tumour growth (tumour budding) in tumour tiles classified as imCMS4 from all three cohorts. imCMS1 tiles more frequently contain lymphocytic infiltration and focal mucinous differentiation, although this feature was less frequent than previously observed in CRC resection specimens (compare TCGA, bottom (Figure 6)), which is consistent with a lower representation of mucinous differentiation in RC. imCMS2 and imCMS3 features were consistent with previously described patterns, with imCMS2 showing epithelial-rich glandular and cribriform tumour growth with focal comedo-like necrosis while imCMS3 was characterized by glandular differentiation with tubular growth, focal mucin and a minor villiform component. Bioinformatic deconvolution of cell composition supported the observed tile-level associations. In biopsy specimens classified as imCMS4, a significantly higher frequency of fibroblast signatures were observed, while imCMS1 samples showed a tendency towards increased immune signatures, supporting biological interpretability of imCMS predictions at the case level.

**Figure 6.**
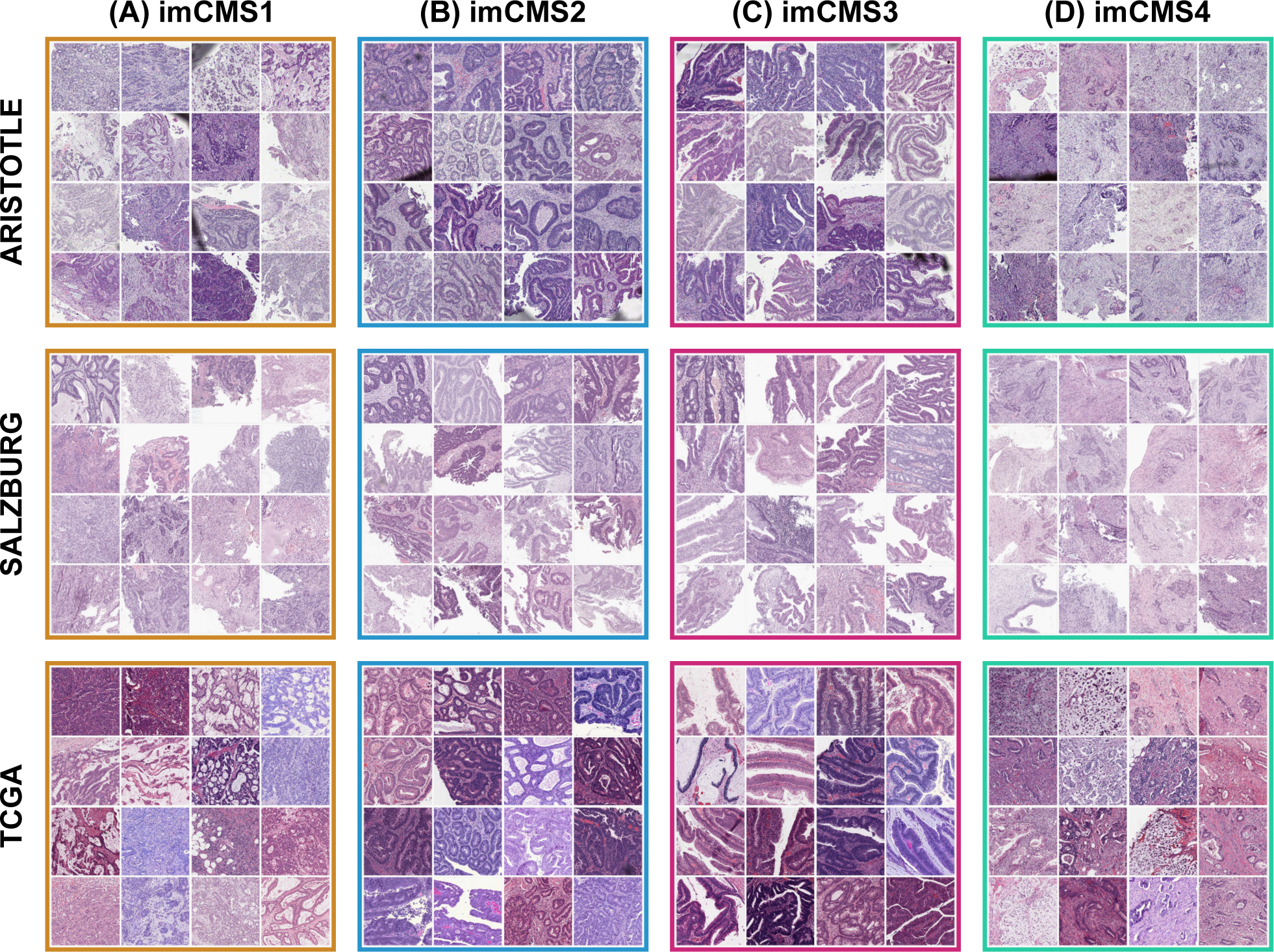
Gallery of image patches (side 630*µ*m) from three cohorts (ARISTOTLE, SALZBURG, TCGA). For each cohort, we selected the 16 image patches from different patients, with highest probability score for each imCMS class (based on the ensemble model of experiment [E3a]). Visual interpretation confirms the existence of distinct morphological patterns within each predicted imCMS class. (A), imCMS1: increased lymphocytic infiltrates, poor tumor differentiation, focal mucin; (B), imCMS2: glandular differentiation with cribriform growth patterns and comedo-like necrosis associated with imCMS2; (C), imCMS3: glandular differentiation with ectatic mucin-filled glandular structures in combination with a minor component showing papillary and cribriform morphology; (D), imCMS4: prominent desmoplastic stromal reaction and dissociative tumor growth (tumor budding).

## DISCUSSION

There are currently no clinically established predictive markers for response to neoadjuvant chemoradiotherapy in RC. Decision making for RC patients and strategies for assignment to neoadjuvant treatment protocols are still taken at the cohort level and it is challenging to predict which patients will respond to neoadjuvant chemoradiotherapy. Current pathology assessment of pre-operative biopsies is limited to the confirmation of cancer diagnosis and a limited panel of molecular studies such as testing for mismatch-repair deficiency (MMRd) and the testing for common driver mutations if clinically desired. Recent studies have demonstrated a strong benefit of neoadjuvant treatment of patients with MMRd RC with PD-1 Blockade, but with a frequency of approximately 2-3%, this genotype is infrequent in RC [22]. Other studies have suggested that the absence of tumour budding [23, 24], low stromal content [24] or the quantification of cytotoxic T-cells [25] may aid in identifying patients with favourable prognosis, but these methods are based on subjective visual features and incompletely capture the complex biology related to neoadjuvant chemoradiotherapy response. Better methods to supply a biologically informed and clinically relevant classification of RC biology from pre-operative biopsy material are therefore needed. Consensus molecular subtyping may aid transcriptome-based staging in both colon and rectal cancer.

Based on their distinct biology and clinical behavior, CMS1 and CMS4 subgroups play a key role for prognostication and may support the development and assignment of patients to biologically informed precision treatments [11, 12]. Specifically, the CMS1 subgroup (immune subtype) is highly enriched for MSI cases and the CpG Island Methylathion (CIMP-high) subgroup of colorectal cancers. Gene enrichment analysis has shown high expression of immune response genes, in particular interferon signaling as well as the wound healing signature[11]. Analysis of the tumor microenvironment has demonstrated dense infiltration by anti-tumoral immune populations, in particular CD8+ cytotoxic T-lymphocytes, which play a key role in mediating the anti-tumoral treatment effect of radiotherapy. These biological features support an improved prognosis, increased propensity for response to radiotherapy and may indicate an increased likelihood of response to immune checkpoint inhibitors. In contrast, CMS4 (mesenchymal subtype) comprises stroma-rich tumors with an activation of TGF-*β* signaling pathways, evidence of epithelial-mesenchymal transition and invasive growth pattern [11, 12]. These cases are characterized by a poor prognosis and increased resistance to radiotherapy treatment as well as conventional chemotherapy protocols [26]. Studies have demonstrated an improved response and prolonged survival of CMS4 patients to irinotecan as compared with oxaliplatin-based chemotherapy, but overall improvements were limited [26]. Novel treatments will be needed (in particular TGF-*β*-targeting anticancer agents) for subtype-specific interventions in CMS4 and to further improve outcomes.

In an earlier study [13], we developed an image-based approach to predict the consensus molecular subtypes from WSIs of clinical CRC specimens and provided the proof of principle for generating imagebased molecular calls even with minimum input material. In a clinical setting, such image-based morphomolecular classifiers have the benefit to offer a level of interpretability linked to known molecular profiles, as opposed to black-box classifiers whose interpretation is limited. The predictive ability of image-based molecular calls for response to chemoradiotherapy remained an open question. For the current study, we re-implemented the imCMS analysis pipeline and trained classifiers using a combination of CRC resection and biopsy cohorts, across different stages. We found that combining these datasets was a viable option to enable generalization to external biopsy cohorts without having to rely on domain adversarial training or fine-tuning as previously required [13]. This incremental improvement showed that such computational pipelines can directly and accurately classify images in new cohorts without the need for additional data or re-training. Despite distributional shifts between training cohorts (e.g., biopsy sampling artefacts, relative proportions of different tissue morphology) can make models subject to learning cohort-specific features that can limit classification performance, we observed a consistent improvement of performance when combining resection and biopsy modalities for training. This result is the most striking when comparing the performances of the models tested using TCGA: models trained solely with biopsy data (GRAMPIAN or ARISTOTLE) were not able to generalize unless resection data (FOCUS and SPINAL) was used. Furthermore, we conjecture that combining these datasets enables learning of modality-agnostic features that are invariant to inter-cohort variations. This hypothesis is supported by our visual assessment of the image patches with highest probability scores from different patients across the TCGA, ARISTOTLE and SALZBURG cohorts, which illustrates the stability of morphology of top-contributing tiles across independent cohorts. Yet, this visual assessment and the suggested associations observed between local morphology and CMS classes should be further studied in bottom-up approaches and further confirmed in conjunction with the establishment of the CMS classes in small fields of view.

The significant associations between imCMS1 and pCR to neoadjuvant LCRT and between imCMS4 and lack of pCR to neoadjuvant LCRT suggest the existence of predictive morphological features that our models were able to capture. This result paves the way for future work for the validation of such a tool as a support for clinical decision in the neoadjuvant treatment setting. Specifically, imCMS1 calls could identify patients for total neoadjuvant treatment with favorable prognosis. Due to an enrichment for immune-activated cases and MMRd cases, imCMS1 cases could also have a greater likelihood to achieve complete remission with immune-checkpoint blockade [27, 28], but this association requires further study. In contrast, patients classified as imCMS4 could be selected for clinical trials adding additional chemotherapy or biological agents to CRT, as the likelihood for response to standard LCRT protocols with single agent fluoropyrimidine is low. This result is aligned with current understanding of CMS4-associated biology: based on the high stromal content and TGF-*β* signatures in this subset, resistance to cytotoxic treatment is common [29]. For imCMS4 patients, tumour-stroma-based therapeutic targeting may therefore offer a potentially efficacious and biologically informed treatment alternative [27, 28]. Ten Hoorn et al. suggest that prospective studies are required to further establish the CMS taxonomy and confirm treatment efficacy by CMS subtype in clinical practice [12]. However, CRC subtyping via current RNA sequencing technologies is deficient, in scenarios with low tumour material: other modalities such as biopsy imaging thus constitute a promising, fast and cost-effective alternative. Using an image-based approach, we demonstrate successful imCMS classification for all samples in the present study, compared to technical failure rates of up to 35% using state-of-the-art panel sequencing approaches [13].

Current ESGE guidelines [19] recommend sampling a minimum of six biopsies for the diagnosis of colorectal carcinomas, to ensure sampling of tumour fragments, and to reliably represent the overall tumour phenotype. Yet whether this number of biopsies is optimal to determine the classification of CRC according to biological subtypes remains an open question. We thus undertook comprehensive simulation experiments on existing fully digitized clinical cohorts to capture the morphology related to transcriptional CMS calls and to address whether clinically established sampling protocols are sufficient to describe tumour heterogeneity at the gene-expression level. As expected and due to the spatially heterogeneous nature of CRC tumours [30, 31], our results suggest that sampling less than five tumour biopsies is not sufficient to properly predict the CMS call that would be obtained from an equivalent resection specimen of the same tumour. Our results corroborate the 2021 ESGE recommendation as our experiments show on two independent external test datasets of 147 and 266 patients, that five or more standard tumorous biopsy fragments are sufficient to reliably capture the global tumour phenotype needed to achieve CMS classification performance with fidelity close to full resection specimens. Thus, the current sampling protocol established in clinical practice is likely sufficient to capture tumour biology from several cancerous fragments, and to enable informed patient stratification by computational analysis.

A limitation of the current imCMS pipeline is the reliance on manual annotations of tumour regions as a pre-processing step, making the whole pipeline semi-automated. We suggest that future work should investigate strategies to either fully automate this step or to accurately classify WSIs independently of localized tumour regions. The pipeline of imCMSv1.5 was designed as an incremental extension of the work of Sirinukunwattana et al. [13], yet with the goal of increasing classification and generalization performance. Although model performance was reported in held out test sets in full transparency, the variation of performance observed across the different test sets suggest the existence of hidden dataset-related factors impacting model classification. The identification of these factors and the improvement of model robustness remain a priority for the development of future versions of imCMS. Other deep learning architectures and training procedures such as recent proposed solutions to biopsy image classification problems [32, 33] and consideration of model uncertainty beyond ensemble majority voting should be done in future work. A weakness of our simulation experiments is the strong assumption for randomness of the sampling distribution of biopsy fragments and we concede that the distribution of true biopsy fragments may be different from our simulation, e.g., tumour fragments from the lumen are more likely to be sampled in a real-world scenario. The effect of these degrees of approximation should be investigated in future work and across cancer types. We also recognize that the guidelines requiring six biopsies per case is designed to ensure there is a high chance of identifying invasive cancer, rather than slough or non-invasive malignancy. Here, our approach shows five or more biopsy fragments containing invasive tumour are required to achieve optimal imCMS classification. Although we did not observe any significant difference in terms of distribution of cell types between the studied resection and biopsy cohorts, such sampling information should be accounted for in future work. Beyond prediction of CMS classes, other molecular signatures of CRC proposed in the literature [34, 35] are relevant candidates to identify associations with treatment outcome.

To conclude, we found that deep learning models can automatically capture the morphology associated with transcriptional CMS in imaged biopsies from unseen cohorts. We found that patients stratified according to biopsy-based imCMS respond differently to neoadjuvant LCRT. The results of this study therefore support the development of an inexpensive clinical tool to assign patients to subtype targeted biological interventions in future clinical trials. Beyond CMS in CRC, the on-going development morpho-molecular classification models across cancer types and molecular signatures offer a new type of cost-effective computational tools to support clinical decision making [18, 36]. This surrogate for transcriptional analysis can also be of use for research studies with restricted funding, or to revisit existing trial cohorts by studying associations between image-based CMS classification and clinical variables of interest without the need for extra tissue material.

## METHODS

### Study Design

The study design, cohorts and aims are outlined in (Figure 1) and detailed methods for all experiments and statistical analysis are provided in (Supplementary Material and Methods).

### Rectal Cancer LCRT Treatment Protocol

All patients from the GRAMPIAN, ARISTOTLE and SALZBURG cohorts included in the study received a “standard” treatment protocol for advanced RC by pelvic irradiation (45-50.4Gy in 25 fractions over 5 weeks) combined with Capecitabine (825mg*·*m*^−^*^2^ BD on treatment days). Detailed information on these cohorts is available in Supplementary Material and Methods. The primary endpoint for all cases was pCR after completion of LCRT as assessed by histopathological analysis of the surgical RC resection specimen by an expert gastrointestinal pathologist according to established guidelines.

### Processing of Tissue Samples and Slide Scanning

For the four S:CORT cohorts, serial 5-*µ*m sections were cut from one pathologist-selected representative tumour block for H&E staining followed by up to nine unstained sections for RNA extraction. H&E slides were reviewed by an expert gastrointestinal pathologist and invasive cancer regions were annotated to guide RNA and DNA extraction by macrodissection. Regions of extensive necrosis and non-tumour tissue were excluded according to standard practice for molecular tumour profiling. All H&E slides were scanned on an Aperio scanner at magnification 20*×* (0.5*µ*m*·*px*^−^*^1^). All digital slides were re-reviewed and a board-certified pathologist annotated tumour regions while excluding areas containing folds or debris. The data filtering procedure, cohort sizes and additional details for each cohort are summarized in (Supplemental Figure S1).

### Transcriptional CMS Classification

For the S:CORT cohorts, RNA expression was obtained by microarray (Xcel, Affymetrix), and raw CEL files underwent the robust multiarray average normalization with the Affymetrix package (v1.56.0) [37] in R [38]. Batch-corrected transcriptional CMS calls were derived for each sample with CMSclassifier using the protocol described in Supplementary Material and Methods. Any variation in the number of patients and slides in the same cohorts used in the study by Sirinukunwattana et al. [13] stems from the updated procedure for transcriptional CMS. For TCGA [39], the same transcriptional CMS calls were used as previously reported by Sirinukunwattana et al. [13].

### Transcriptomic Immune Profiling

The batch-corrected version of the S:CORT transcriptome was used to derive estimates of immune and stromal cell types with MCPcounter [20] and Xcell [21] by applying the original R packages. Although scores for both signatures are not comparable between cell types, they were scaled from 0 to 1 to facilitate visualization.

### Deep Learning imCMS Classification

To develop a WSI-based CMS classifier, we considered the model proposed by Sirinukunwattana et al. [13] as a baseline (imCMSv1), and re-implemented a new version (imCMS v1.5) that facilitates the training procedure, and reproducibility of our experiments while keeping high performance (test macroaverage AUROC in held-out TCGA of .813) and same capabilities for the interpretation of classification results.

The version 1.5 of imCMS is based on the three-stage process illustrated in (Figure 1D). First, manually annotated tumour regions of an input WSI are tiled with patches of size 318*×*318px at magnification 5*×* (*∼*2*µ*m*·*px*^−^*^1^) with 50% overlap, and all tiles with less than 50% overlap with the annotated tumour regions were excluded. Then, all the extracted patches are fed as input to a trained deep learning model that outputs probability scores for each target CMS class. Third, all tile-level probability scores are averaged to produce slide-level probability scores, and the class with the highest score is considered as the imCMS call prediction for the input WSI.

We kept the first stage identical to imCMSv1 but opted for a fixed magnification for tile extraction at magnification 5*×* based on the results of [13], suggesting optimal performance in both resection and biopsy images. With the last two stages, we moved from a count-based assumption to a collective assumption for determination of imCMS calls [40], thus enabling more fine-grained contributions of each tile to the slide-level predictions. Further, for imCMS1.5, we changed the pre-trained InceptionV3 backbone architecture used in imCMSv1 by a randomly initialized customized ResNet architecture, to ensure that our approach can be re-implemented without having to rely on a pre-training procedure. See Supplementary Material and Methods for more details about the implemented model architecture and training procedure.

## DATA AVAILABILITY STATEMENT

The datasets generated during and/or analysed during the current study are available from the corresponding authors upon reasonable request in accordance with the S:CORT data access policy. The TCGA datasets and images analysed in this study are openly and publicly available at: https://portal.gdc.cancer.gov/.

## CODE AVAILABILITY STATEMENT

The source code of the underlying (trained) models is not available due to proprietary reasons.

## ACKNOWLEDGMENTS

The authors thank Aurelien de Reynies for advice on CMS calling in FFPE blocks, Claire Butler and Michael Youdell for excellent managing in S:CORT and the MRC Clinical Trials Unit who provided the clinical data from the FOCUS trial with permission from the FOCUS trial steering group. The S:CORT consortium is a Medical Research Council stratified medicine consortium jointly funded by the MRC and CRUK (MR/M016587/1). The ARISTOTLE trial was funded by Cancer Research UK (CRUK/08/032). This work was supported by the National Institute for Health Research (NIHR) Oxford Biomedical Research Centre. This work was supported by the Research Fund of the Paracelsus Medical University Salzburg, Austria (PMU-FFF R-17/03/090-HUW). Computation used the CTP Lab core resources at the University of Zurich and the Oxford Biomedical Research Computing (BMRC) facility, a joint development between the Wellcome Centre for Human Genetics and the Big Data Institute supported by Health Data Research UK and the NIHR Oxford Biomedical Research Centre. RW is supported through the EPSRC Center for Doctoral Training in Health Data Science (EP/S02428X/1), Oxford CRUK Cancer Centre. JR is supported through the NIHR Oxford Biomedical Research Centre, the Oxford CRUK Cancer Center, and holds an adjunct appointment at the Ludwig Institute of Cancer Research at the University of Oxford. TM gratefully acknowledges funding by the Medical Research Council and Cancer Research UK. VHK gratefully acknowledges funding by the Swiss National Science Foundation (P2SKP3_168322/1 and P2SKP3_168322/2), and the Promedica Foundation (F-87701-41-01). The results published or shown here are based in part upon data generated by the TCGA Research Network established by the NCI and NHGRI. Information about TCGA and the investigators and institutions who constitute the TCGA research network can be found at http://cancergenome.nih.gov. The funders played no role in the analyses performed or the results presented. The views expressed are those of the author(s) and not necessarily those of the NHS, the NIHR or the Department of Health.

## AUTHOR CONTRIBUTIONS

MWL, ED, KS, TM, JR and VHK jointly conceived the study; MWL, ED, KS, TM, JR and VHK designed the study; MWL, ED, KS, TM, JR and VHK drafted the manuscript; MWL developed and analysed deep learning models; ED performed bioinformatic and statistical analysis; MWL, ED, VHK performed data interpretation and analysed the experiments; LS, GM, SDR, AB, DSM, SG, EK, DN, FH, RG, PD, PQ, LW, VHK, TM obtained and categorised clinicopathological and molecular data; KS, RW, JR, LS, GM, SDR, AB, DSM, SG, EK, DN, FH, RG, PD, PQ, LW, VHK, TM provided important intellectual input, provided critical resources or funding, and critically reviewed the study design; All authors have read and given approval of the final manuscript.

## SUPPLEMENTARY INFORMATION

**Figure S1.**
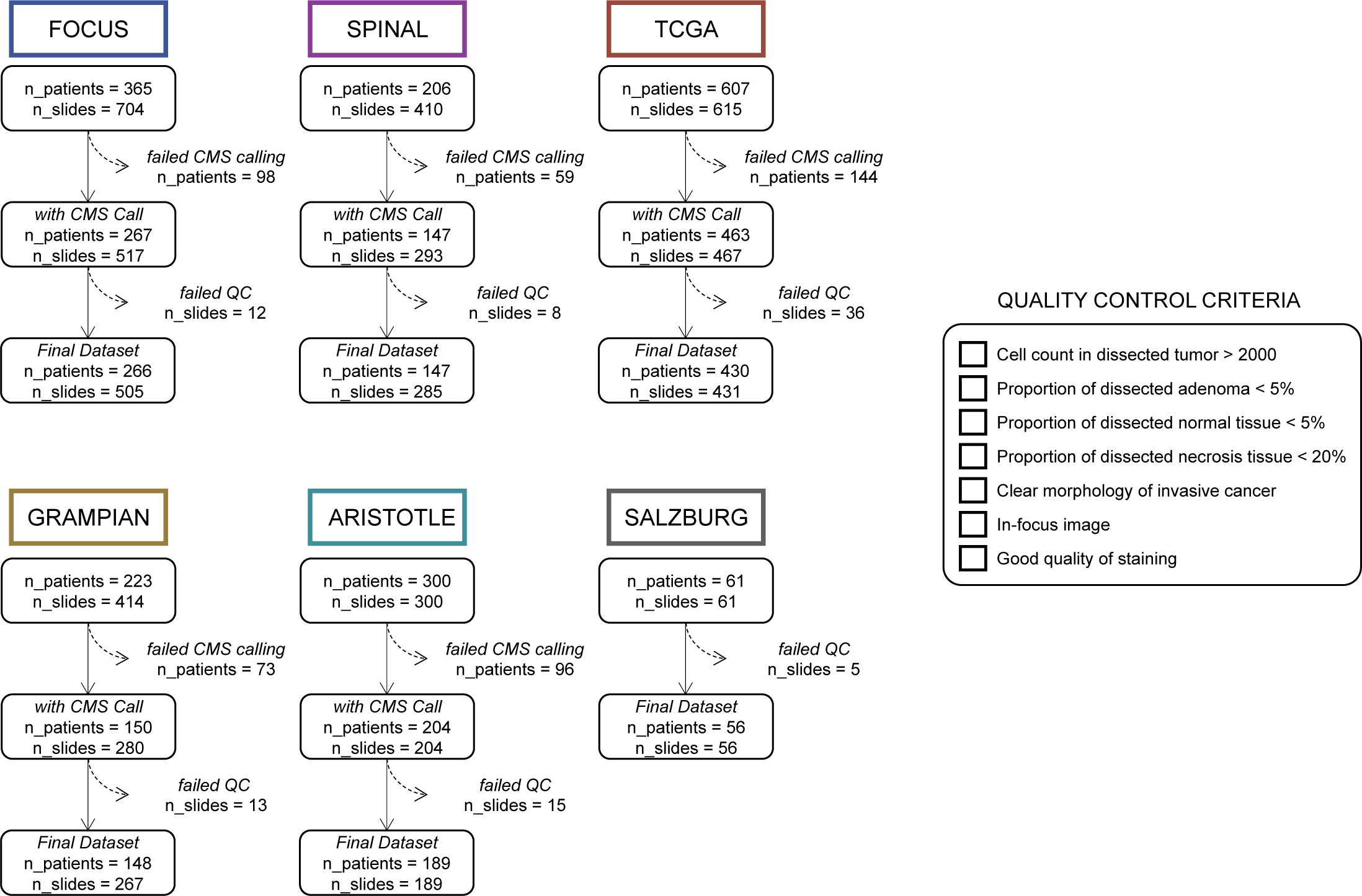
Detailed description of the six cohorts used in this study. For each cohort, a flowchart indicates how many patients and images were successively excluded from the study based on the failure of transcriptional CMS calling, and based on visual assessment of the quality of the WSIs using the criteria listed on the right.

**Figure S2.**
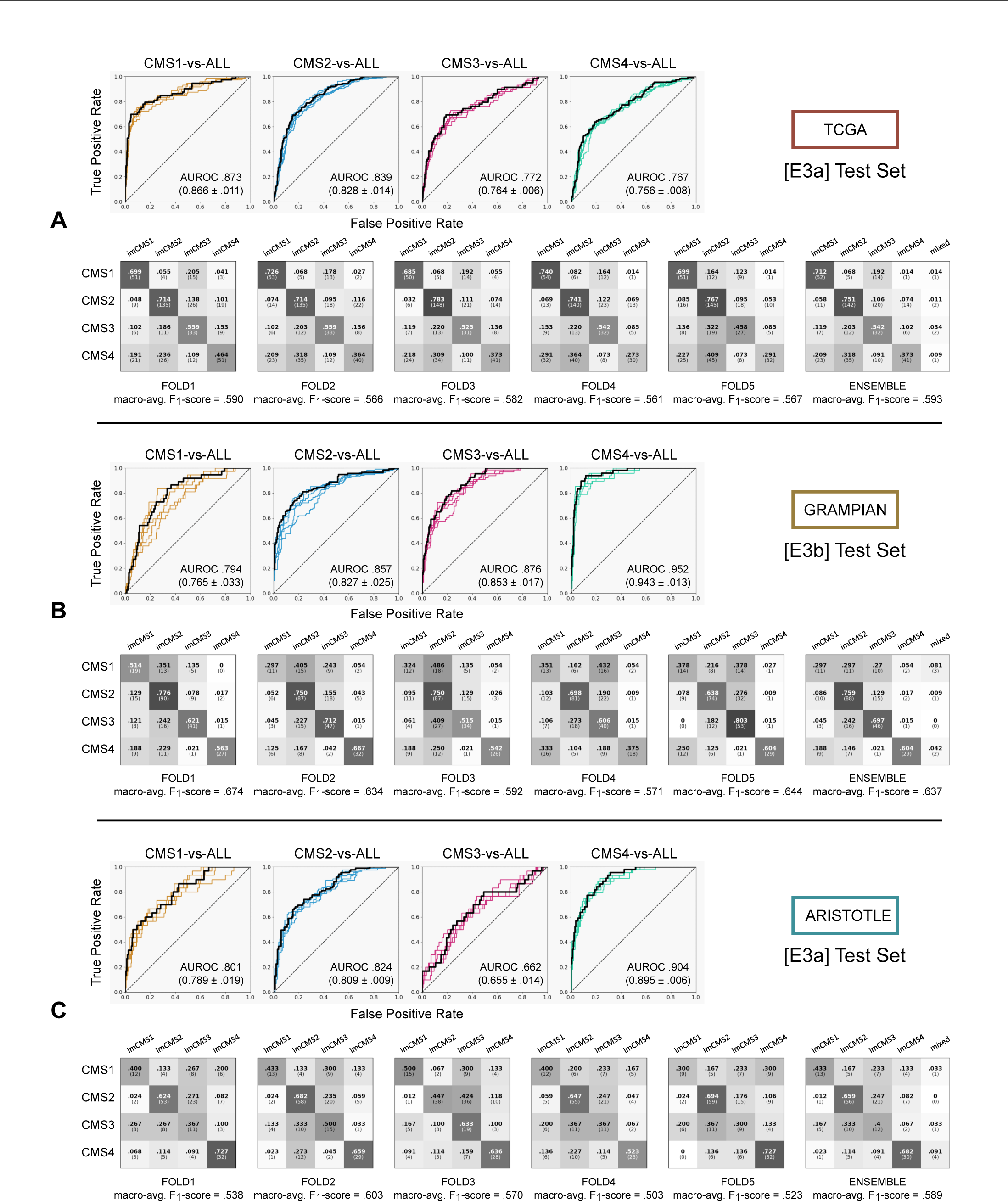
Detailed Receiver Operating Curves (ROC) and confusion matrices (CMs) of the trained imCMS models of experiments [E3a] with TCGA (A) and ARISTOLE (C) as test sets, and of experiments [E3b] with GRAMPIAN (B) as test set. Area under the ROC (AUROC) are shown for each CMS class and each of the five trained models of each experiment. Results of majority voting of five models (ensemble model) are reported in the right-most CMs. Cases without absolute majority were classified as “mixed”. The reported macro-average F1-scores are derived from the confusion matrices and are ranging from 0 to 1, with 1 indicating perfect classification.

**Figure S3.**
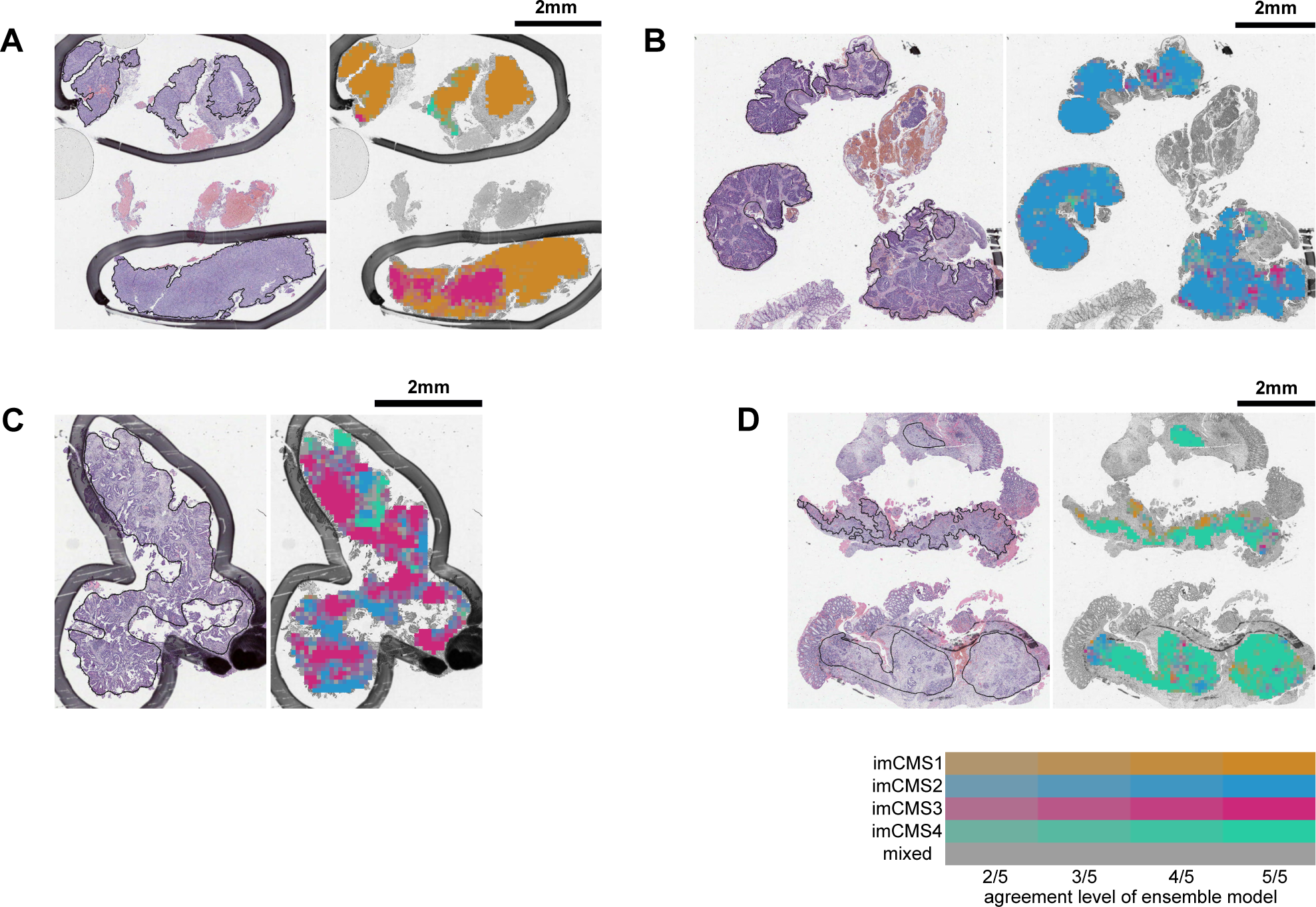
Examples of classification maps generated with the ensemble model of experiment [E3a] for correctly classified WSIs of the ARISTOTLE cohort. Pathologist annotation of invasive cancer regions are indicated by black lines (digital) or pen-marks (analogue). Ground-truth transcriptional CMS calls: CMS1 (A); CMS2 (B); CMS3 (C); CMS4 (D). Color visualization is based on the highest voted tile-level predicted imCMS class among the five trained models that form the ensemble model of experiment [E3a] as indicated in the bottom-right legend. Classification maps illustrate the spatially resolved imCMS calls across biopsy samples including samples with some level of pervasive heterogeneity as previously described for both image-based as well as molecular analysis methods [13, 30, 31].

### Additional Details on Clinical Cohorts

To train and validate the deep learning models developed in this study, H&E-stained tissue specimens of diverse stage and clinical settings were used from cohorts from the Medical Research Council (MRC) and Cancer Research UK (CRUK) Stratification in COloRecTal cancer (S:CORT) programme:

FOCUS (colon and rectal resections, n=365 patients; n=704 slides), randomised clinical trial testing different strategies of sequential and combination chemotherapy for patients with advanced CRC after surgical resection (MRC FOCUS, ISRCTN79877428) [41].

SPINAL (colon and rectal resections, n=206 patients; n=410 slides), patients with primary tumors without previous treatment balanced according to T stage, N stage, location (colon/rectum) and recurrence/metastatic disease (either at diagnosis or during follow-up). Samples were obtained from Birmingham and Manchester Hospitals, United Kingdom and the COIN clinical trial (ISRCTN27286448) [42].

GRAMPIAN (rectal preoperative biopsies, n=233 patients; n=414 slides; sequential cohort of high-risk RC with threatened or involved circumferential rectal fascia on pre-treatment MRI scan treated at Aberdeen Royal Infirmary, United Kingdom. We report a median number of fragments containing tumor content across the biopsies of the GRAMPIAN cohort of 4.

ARISTOTLE (rectal preoperative biopsies, n=300 patients; n=300 slides, (base cohort)) UK national clinical trial (ISRCTN09351447) [43] which compared the efficacy of standard CRT with (intervention) or without (control) irinotecan in high-risk RC with threatened or involved circumferential rectal fascia on pre-treatment MRI scan. Cases were selected from the control arm of the ARISTOTLE trial and in whom biopsies were available for molecular analysis. Pathological response was assessed centrally according to a pre-specified pathology protocol using the Dworak method (NPW Leeds). We report a median number of fragments containing tumor content across the biopsies of the ARISTOTLE cohort of 4.

SALZBURG (rectal preoperative biopsies, n=61 patients; n=61 slides); sequential cohort of high-risk RC treated at the III^rd^ Department of Internal Medicine of the Paracelsus Medical University Salzburg, Salzburg, Austria; Patients received neoadjuvant long-course chemoradiotherapy with single agent capecitabine as detailed in “RC Treatment”. Pathological response was assessed by detailed histopathological assessment of the resection specimen, undertaken 6-12 weeks after CRT using the Dworak method.

The ARISTOTLE and SALZBURG cohorts were strictly selected to have undergone the same treatment protocol for advanced rectal cancer by pelvic irradiation combined with single agent fluoropyrimidine.

Clinical data was anonymized by S:CORT number and was provided comprising demographic data, baseline stage generated from pre-treatment pelvic MRI scans and CT scans TAP, and outcome data.

TCGA H&E-stained tissue samples of The Cancer Genome Atlas Colon Adenocarcinoma (TCGA-COAD) and Rectal Cancer Rectum Adenocarcinoma (TCGA-READ) data collection (colon and rectal resections, n=430 patients; n=431 slides) described and made available by the TCGA Research Network at https://www.cancer.gov/tcga [39].

#### Ethical Approval

All samples in the S:CORT cohorts were obtained following individual informed consent and ethical approval by the National Research Ethics Service in the United Kingdom (ref 15/EE/0241; IRAS reference 169363). The SALZBURG cohort was reviewed by the ethical board of the provincial government of Salzburg, Austria (415-E/2343/5-2018), although under Austrian law informed consent is not needed for research use and is therefore not available for all cases.

#### Transcriptional CMS classification

To obtain robust CMS calls, initially three different CMS classifications were derived from three different transcriptomic datasets. The first transcriptome was composed of each single cohort analyzed independently, the second from a homogenized transcriptomic dataset correcting batch effects by cohort with ComBat and the third from a dataset including all S:CORT cases also batch-corrected by cohort. For the three transcriptomes, gene-level data was obtained with the mean of probe sets linked to each NCBI Entrez Gene ID according to the latest xcel annotation file (v36). Then, CMS calls were derived with the random forest CMSclassifier with the default posterior probability of 0.5. Random forest classification of FFPE samples leads to an increased frequency of unclassified samples compared to published datasets derived from fresh frozen material [13].

To derive calls with lower frequency of unclassified cases, we additionally computed single sample predictor calls after row-centering the expression data. CMS calls were generated when there was a match between both methods (RF and single sample predictor without applying any cut-off) as previously [13]. After obtaining three CMS calls from the three transcriptomes for each sample, a final call for training imCMS was computed when all three CMS calls were matching or when two of them were matching and one was unclassified. This method ensures high quality, robust transcriptomic CMS calls from FFPE tissue.

#### Deep Learning Model architecture and Training Procedure

The imCMS v1.5 classification model is based on a customized ResNet convolutional neural network [44] with 55 layers that make use of rotation-equivariant convolutional layers as described in [45]. This modification provides guarantees that the model predictions do not depend on the orientation of the input (for 90-degree rotation angles), which was shown to be beneficial across several classification tasks for histopathology image analysis [45–47]. This architecture was designed to take image patches of size 318*×*318px (*∼*636*×*636 *µ*m^2^ at magnification 5*×*) as input and to output the probability scores (softmax-activated vector of four logit values) for the transcriptional CMS class of the tumor from which image patches originated.

For each training/validation split of the five folds of each investigated multi-cohort development set in this study, we trained a model according to a two-stage procedure. In a first stage, under a weakly supervised framework, each model was trained to predict slide-level CMS classes from tile-level input image patches. Each model was trained via minimization of the cross-entropy loss using random batches of size 64 from the training partition with uniform distribution of transcriptional CMS. Models were trained with gradient descent with initial learning rate 0.01, cycling cosine annealing (period of 10 000 iterations), momentum 0.9 and weight decay 0.0001. During training, data augmentation was used to improve the robustness of the model against appearance variability of histology images (random horizontal flip, random hue rotation, random gamma correction, channel-wise random intensity shift). Over training (100 000 iterations), we saved the state of the model that minimized the cross-entropy in the corresponding validation partition. In a second stage, we froze the 53 first layers of the trained model from the first-stage and fine-tuned the last two layers in order to optimize slide-level classification performance. To this purpose, we pre-computed the vector output of the 53rd layer for all training image patches and re-trained the last two layers by using batches that include all the image patches of 16 random WSIs of the training partition with uniform distribution of transcriptional CMS. During this second training stage, all the predicted probability scores for all the image patches from the same WSI were averaged and the model was optimized via minimization of the cross-entropy loss for the averaged probability scores.

To assess the generalization of imCMS on unseen cohorts, we trained all models with data from a fixed development set, and systematically reported performance with test data from held out cohorts that were not used for training, fine-tuning or any part of the model selection procedure.

#### Statistical Analysis

Four logistic regression models for presence vs. lack of presence of each imCMS class were built for the endpoint of pCR. Models were adjusted by cohort (i.e., ARISTOTLE and SALZBURG) and the clinical confounders pretreatment T stage and pretreatment N stage, determined from pretreatment MRI assessments (i.e., at the time of clinical decision). The category ‘mixed imCMS’ was not analysed as it only contained two cases. p-values <0.05 were considered statistically significant. Statistical analyses were conducted using R [38].

